# Semantic Segmentation to Extract Coronary Arteries in Invasive Coronary Angiograms

**DOI:** 10.1101/2020.05.26.20103440

**Authors:** Chen Zhao, Robert Bober, Haipeng Tang, Jinshan Tang, Minghao Dong, Chaoyang Zhang, Zhuo He, Yu-Ping Wang, Hong-Wen Deng, Michele L. Esposito, Zhihui Xu, Weihua Zhou

## Abstract

Coronary artery disease (CAD) is the leading cause of death worldwide, constituting more than one-fourth of global mortalities every year. Accurate semantic segmentation of each artery using invasive coronary angiography (ICA) is important for stenosis assessment and CAD diagnosis. However, due to the morphological similarity among different types of arteries, it is challenging for deep-learning-based models to generate semantic segmentation with an end-to-end approach. In this paper, we propose a multi-step semantic segmentation algorithm based on the analysis of arterial segments extracted from ICAs. The proposed algorithm firstly extracts the entire arterial binary mask (binary vascular tree) using a deep learning-based method. Then we extract the centerline of the binary vascular tree and separate it into different arterial segments. Finally, by extracting the underlying arterial topology, position and pixel features, we construct a powerful coronary artery segment classifier based on support vector machine. Each arterial segment is classified into left coronary artery (LCA), left anterior descending (LAD) and other types of arterial segments. We tested the proposed method on a dataset with 225 ICAs and achieved artery classification accuracy of 70.33%. The experimental results show the effectiveness of the proposed algorithm, which provides impressive performance for analyzing the individual arteries in ICAs.

## 1. Introduction

Coronary artery disease (CAD) is the leading cause of morbidity and mortality in the United States and costs about 350 billion dollars annually [1]. Invasive coronary angiography (ICA) remains the gold standard for the diagnosis of CAD [2]. ICA involves injection of contrast media into the epicardial arteries with acquisition of continuous fluoroscopy. Detection of CAD is performed by visually comparing diseased arterial segments to normal arterial segments and is essential for the diagnosis and treatment.

Semantic segmentation of coronary vessels is extremely important for clinical decisions regarding mechanical revascularization. For these clinical decisions, automatic identification of correct anatomical branches provide meaningful information for automatic diagnosis report generation and region of interest visualization [3]. Meanwhile, successfully detecting the percent stenosis of a coronary artery improves the diagnostic efficiency and confidence, and also influences management [4]. However, due to the overlap of arteries on ICA and the inter-subject variation of coronary artery segments, identification of the arterial branch is challenging. According to the prior knowledge [5], the left main coronary artery (LMA) is the artery that arises from the aorta above the left cusp of the aortic valve and perfuses the anterior, septal and lateral walls of the left ventricle. The LMA branches into the left anterior descending artery (LAD), which courses between the left and right ventricles towards the apex along the anterior interventricular sulcus, and the left circumflex artery (LCX) which courses laterally along the atrioventricular groove. Typically, practioners analyze the entire vascular tree according to the position and morphology of LMA, LAD and LCX.

Recently, deep learning techniques, especially variants based on convolutional neural networks (CNN), have been employed on coronary artery segmentation in ICAs. Nasr-Esfahani et al [6], implemented a CNN network which contains two convolutional layers to predict the patched arterial binary mask. The same group also used a CNN to classify the central pixel within the cropped patches into arterial pixel or background [7]. The AngioNet employed DeepLabV3+ network as the backend for by coronary artery segmentation and achieved a Dice score of 0.864 [8]. Our recent published network [9], feature pyramid U-Net++, was developed based on U-Net++ and integrated the feature pyramids to capture feature maps from different scales; the proposed network achieved a Dice score of 0.8899 on our dataset with 314 annotated coronary arteries. Even though these proposed approaches have achieved a state-of-the-art performance on artery segmentation in ICA images, the methods are not suitable for semantic segmentation to extract individual vessel segments. The existing approaches extracted the entire vascular tree in ICAs and did not take the position and topology of vessel segments into consideration. Hence, current approaches have a major limitation: it is difficult to assess arterial anatomy in ICAs due to the very low contrast, moving objects, inconstant contrast agent, and limited view angles [10]. In addition, due to the morphological similarity of different types of arteries, it is challenging for pixel-intensity-based models to discern each arterial segment and generate semantic segmentation.

The topology is an important factor in arterial identification, which inspires us to convert arteries and their connections into graphs. The non-traditional features, such as the node degrees, can be combined with the pixel-derived features in the semantic segmentation. The problem of semantic segmentation here will be converted into a problem that classifies the type of an unlabeled arterial segment. Figure 2 illustrates our overall workflow. In this paper, we propose a new machine learning-based approach to extract individual coronary arteries from ICAs by incorporating global position, topology and pixel information. The focus of this work is on the classification of the major and branch arterial segments into LMA, LAD, LCX, diagonal branch (D1), obtuse marginal branch (OM1), and ramus intermedius branch (RI). We first extract the entire vascular tree from the ICA images by using our previously designed feature pyramid U-Net++ (FP-U-Net++) [9]. After that, we generate the centerline of the whole vascular tree, and find the key points to generate the vessel graph. Finally, we extract positional and topological features from vessel segments and perform the semantic segmentation.

**Fig. 1.**
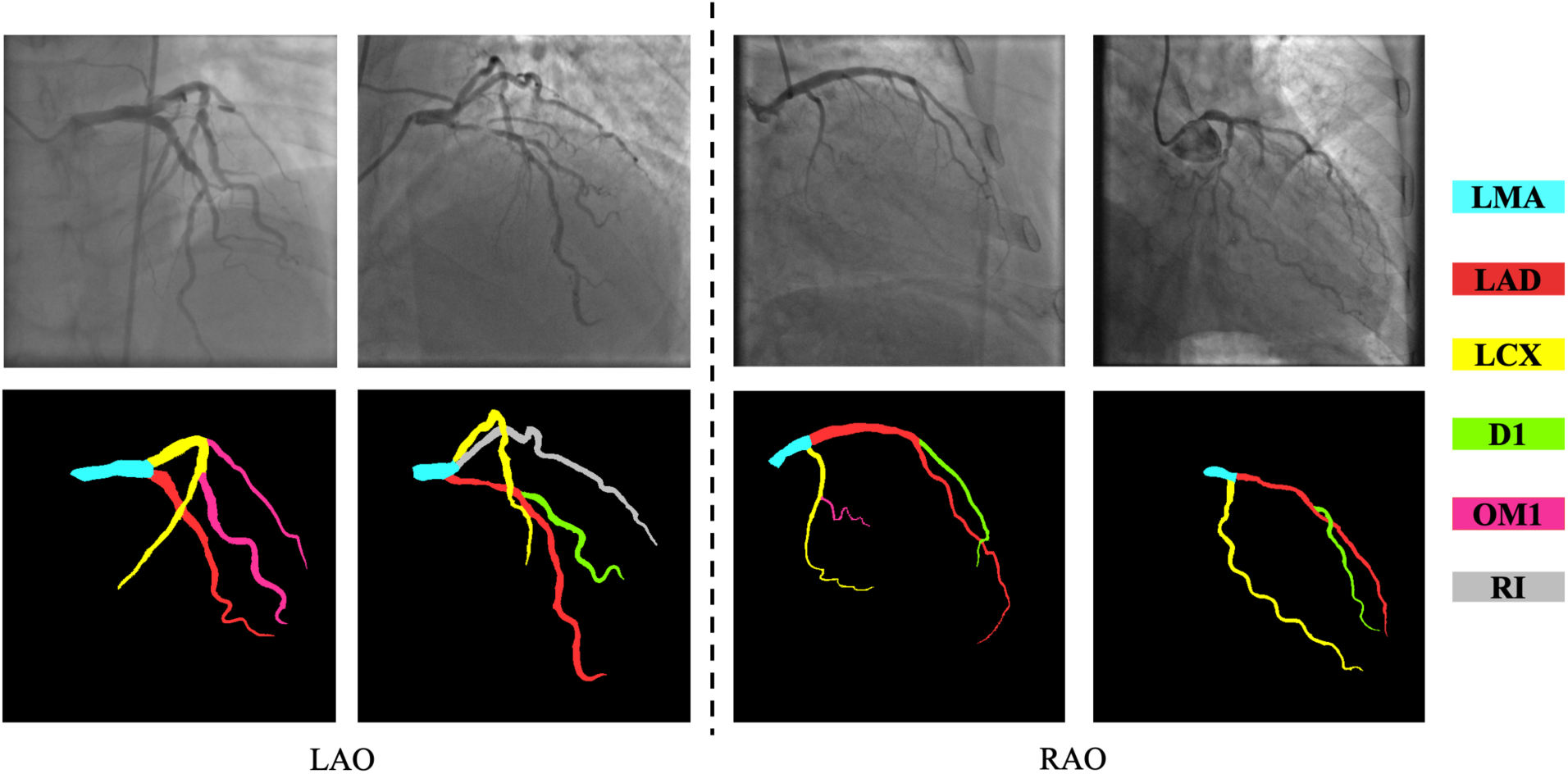
Illustration of semantic segmentation of arteries in (a) Left Anterior Oblique (LAO) view, and (b) Right Anterior Oblique (RAO) view. For each view, the raw image and semantic maps are juxtaposed vertically. Arteries with different colors in the semantic maps represent different types of arteries.

**Fig. 2.**
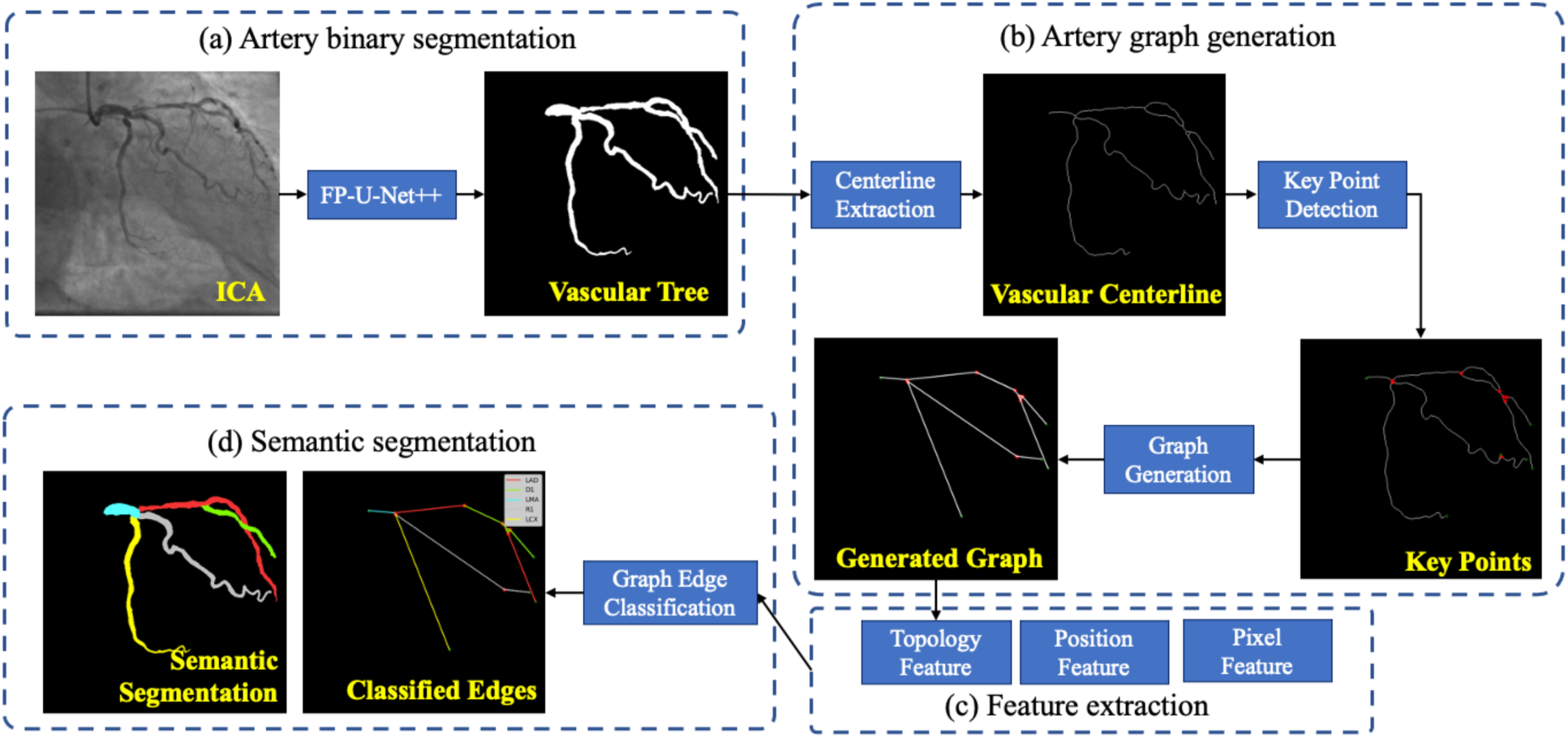
Workflow of coronary artery semantic segmentation. FP-U-Net++ is a deep learning based neural network for coronary artery segmentation, which was published by Zhao et al. in [9].

## 2. Materials and Methodology

### 2.1. Enrolled subjects

This retrospective study enrolled 113 patients who received ICA from February 26, 2019 to July 18, 2019. It was approved by the ethics committee of The First Affiliated Hospital of Nanjing Medical University. ICA was performed using an interventional angiography system (AXIOM-Artis, Siemens, Munich) and was acquired at 15 frames/sec at the Jiangsu Province People’s Hospital, China. The images sizes of ICA videos were range from 512×512 to 864×864, and the pixel spacing ranges from 0.2 mm to 0.39 mm.

The dataset consists of 225 left ventricle ICAs with 135 LAO images and 90 RAO images. For each patient, at most 3 standard views were selected, and then only one frame was selected from the view video for semantic segmentation. The vessel contours were manually drawn by well-trained operators, confirmed by an experienced interventional cardiologist and then provided to this study as the ground truth. For each angiogram, we annotated LMA, LAD, LCX, D1, OM1, and RI arteries.

### 2.2. Artery tree segmentation

In general, current deep-learning-based models for medical image segmentation are variants of the encoder-decoder based architecture, such as U-Net [11]. Many recent networks employed the classification network with the pre-trained weights in ImageNet [12] as the backbone of the encoder. In U-Net++, the skip-connections are modified by using nested and dense connections [13]. In our latest paper [9],the multi-scale technique is improved by feature pyramids, which are built upon image pyramids and forms a fundamental solution for utilizing features in different scales. To leverage the pyramid features of the hierarchy decoder in U-Net++, we resize the feature maps extracted from different layers and integrate them to generate the final feature map. By using pyramid features, the multi-scale problems are significantly resolved. The input of the network is the raw ICA and the output is the binary mask of the entire vascular tree. Our artery tree segmentation model achieved an average Dice score of 0.8899. Results from this binary segmentation model are used to generate the vascular centerline for coronary artery segment separation and semantic labelling.

### 2.3. Artery graph generation

The artery graph generation process includes centerline extraction and segment separation. Centerline extraction aims at removing the redundant pixels in binary mask while preserving the topology and connectivity of the vascular tree. In our implementation, the centerline of vascular tree is generated by using erosion and dilatation operations [14]. The extracted centerline is thus a representation of the vascular tree which contains three types of the nodes [15]: the degree-one nodes are the end of arteries and the degree-two nodes are connecting point; the nodes whose degree is greater or equal to 3 are bifurcation points.

Each pixel in a centerline is iterated, and then the arterial nodes are extracted by finding the bifurcation and the end points. To find the links between the nodes, each pair of points are added into the graph if they are connected adjacently. After that we will remove the degree-two points and corresponding edges from the centerline to generate the undirect graph.

Each arterial segment is represented by a link between two nodes in the generated graph. Semantic segmentation will label the vessels by determining the type of each graph edge and assigning the vessel type to the arterial segment based on the artery pixels between any of the two adjacent nodes [16], the topology information, and the pixels within the arterial segments.

### 2.4. Artery feature extraction and segment label assignment

The basic idea of arterial semantic segmentation is to classify the segments into different classes of arteries. After that, we will assign the arteries with different class labels to achieve semantic segmentation. As demonstrated in section 2.2, each artery segment is represented by a link between two nodes. When we classify a specific segment, we use the features extracted from this segment and its corresponding edge. For feature extraction from the edge, topology information will be used: the degree of the nodes connected by this edge will be used as the feature. We also extract the position and pixel features from the segment (See table 1). Using the extracted features of each vessel segment, an off-the-shell machine learning classifier is employed to perform artery segment classification and a grid search will be used to find the best classifier (see section 3). By classifying the artery segments into different classes, the semantic segmentation results are generated, and each segment is classified.

**Table 1.** List of features measured for each artery segment

| Type | Index | Feature description |
| --- | --- | --- |
| Pixel feature | 1 | Number of the pixels in the artery segment |
|  | 2 | Length of the centerline in pixels |
|  | 3-6 | Standard deviation, mean, minimum and maximum radius of the artery segment |
|  | 7-8 | Mean and standard deviation of the intensities within the artery segment |
|  | 9-10 | Mean and standard deviation of the intensities within the centerline of the artery segment |
| Position feature | 11-14 | Weighted and absolute centers of the segment positions related to the center of vascular tree |
|  | 15-22 | Weighted and absolute positions of the two key points related to the vascular tree center |
|  | 23-30 | Weighted and absolute positions of the two key points related to the artery segment center |
| Topology feature | 31-32 | Degree of the two key points |
| Projection view | 33 | 1 represents left anterior oblique view, and 0 is right anterior oblique view |

Each artery segment is classified into one of six types, including LMA, LAD, LCX, D1, OM1 and RI. At the training stage, the dataset was split into 70% subjects as a training set and the rest 30% as a test set. As a result, 158 and 67 images were grouped into the training set and test set, respectively. We employ support vector machine (SVM) with kernel functions, random forest, and multiple layer perceptron (MLP) as the classifiers to perform the artery segment classification. All classifiers were implemented by Weka 3.8 [17]. A grid search is performed to find the optimal parameters. The settings of the grid search are shown in Table 2.

**Table 2.** Settings of the grid search for artery.

| Model | Parameter 1 | Parameter 2 |
| --- | --- | --- |
| SVM | Kernel: RBF kernel, polynomial kernel, linear kernel | Regularization parameter: 0.01, 0.1, 1.0, 10, 100 |
| Random forest | Maximum depth: 5, 10, 15, 20 | # Decision tree: [5, 10, 20, 50] |
| MLP | Learning rate: 0.1, 0.01, 0.001 | Layer settings:<br>[20], [20, 10], [20, 20, 10], [30, 30, 20, 10], [30, 20, 20],<br>[30, 20, 20, 10] |

### 2.5. Evaluation metrics

#### Evaluation of artery segment classification

Artery segment classification is a multi-class classification task. The input is the extracted features from the artery segment and the output is the probability distribution of the artery class among 6 types. The class with the maximum probability is regarded as the classification result. We evaluated the performance using accuracy (ACC), sensitivity (SN) and specificity (SP). The SN measures the proportion of the true positive samples that are correctly predicted while the SP indicates the proportion of the true negative samples that are correctly predicted. An ACC, SN or SP of 1 imply a perfect prediction.

#### Evaluation of artery semantic segmentation

Semantic segmentation is to assign labels for each pixel in the image. We evaluated the semantic segmentation performance using ACC and mean intersection over union (mIoU). IoU measures the proportion of the intersection between the segmented pixels and the pixels in the ground truth over the number of pixels in either the segmented pixels or the ground truth. The definition of mIoU is shown in Eq. 1.

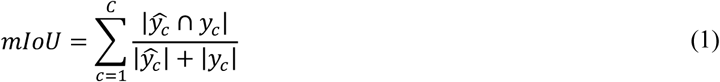

where *C* is the number of classes in the dataset, *ŷ*_*c*_ represents the predicted arterial mask for class *c*, and *y*_*c*_ indicates the ground truth of the coronary artery.

## 3. Experimental Results

### 3.1. Results of artery graph generation

We first applied our binary artery segmentation model [9] to our dataset to generate coronary artery binary mask. Then we extracted the centerlines and generated a graph for each ICA. To obtain the topology structure and separate the vascular tree into different arterial segments, the edge linking algorithm was applied [18]. In Fig. 3 (c), the bifurcation points and end points are annotated by red star and green plus, respectively. By splitting the vascular tree according to the detected key points, the coronary artery segments were obtained. In clinical practice, cardiologists only focus on the main arterial branches, and the branches with limited lengths would be ignored. In our implementation, if the length of the centerline is less than 10 pixels, then it was removed. As a result, in the training set and the test set, 1609 and 729 segments were extracted respectively.

**Fig. 3.**
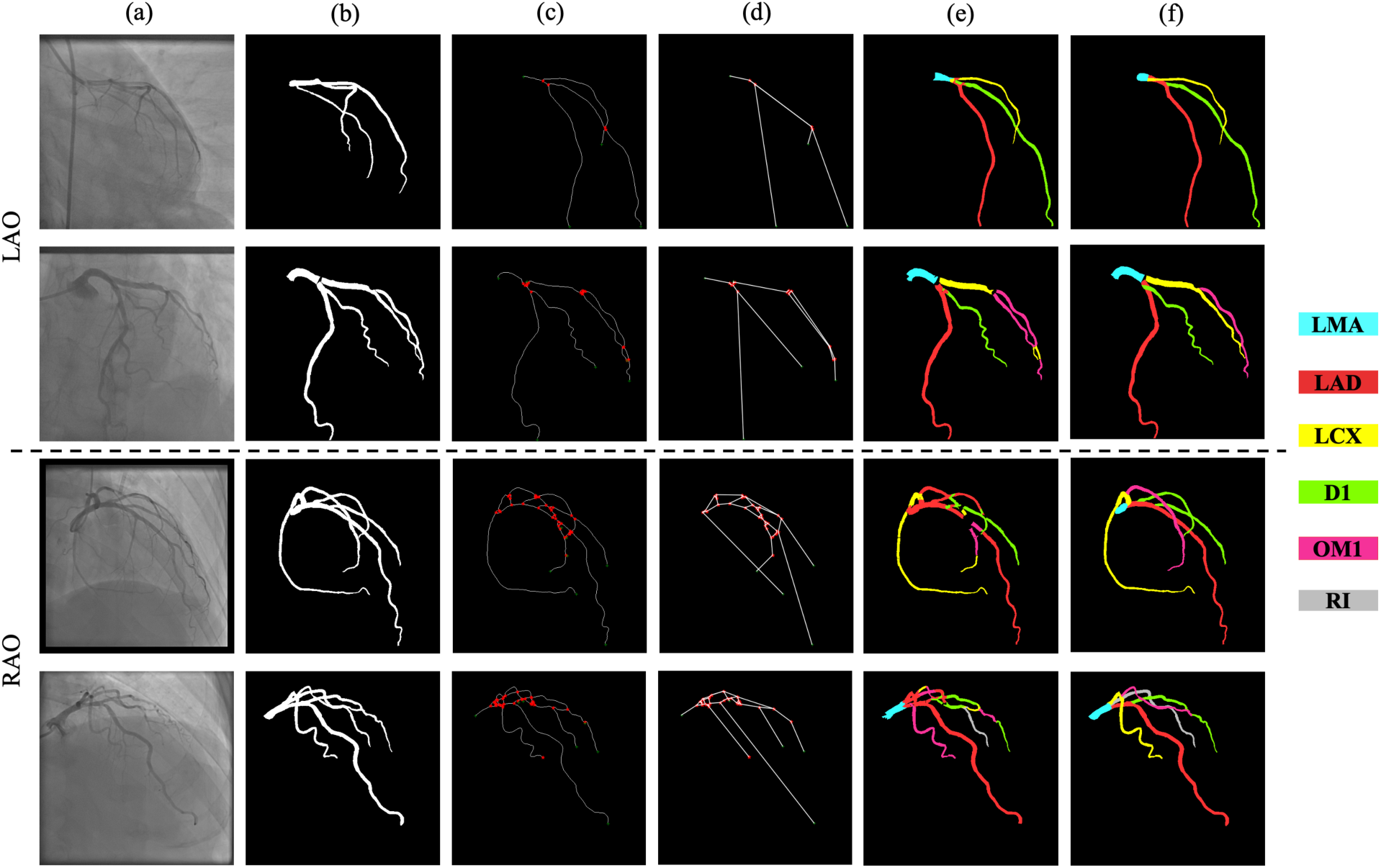
Experimental results on LAO (top) and RAO (bottom) subsets of (a) original ICAs; (b) artery binary segmentation masks; (c) generated coronary centerline and detected key points, where red points indicate bifurcation points and green points are end points; (d) generated artery graph for topological feature extraction; (e) semantic segmentation predictions; and (f) semantic segmentation ground truth. LMA, left marginal artery; LAD, left anterior descending; LCX, left circumflex artery; D1, diagonal branch one; OM1, obtuse marginal branch one; RI, ramus intermedius branch.

### 3.2. Results of artery semantic segmentation

The features extracted from the artery segments were scaled using z-score normalization. We performed the grid search shown in Table 2. The arterial segments were used for model training and the performance was reported according to the prediction on test set. The random forest achieved its best performance with 20 decision trees for ensemble, and the depth for each tree was 5; for MLP, the model achieved the best performance with a learning rate of 0.1 and a layer setting of 20, which indicated that there was only one layer with 20 neurons; for the SVM, the model achieved the best performance with a regulation parameter of 10.0 and a RBF kernel function. The performance of the best classifiers is shown in Table 3. We also evaluated the results using image semantic segmentation metrics. The performance is reported in Table 4.

**Table 3.** Grid search results for coronary artery segment classification.

| Model | ACC | SN | SP |
| --- | --- | --- | --- |
| RF | 0.6498 | 0.6077 | 0.5929 |
| MLP | 0.6475 | 0.5982 | 0.5789 |
| SVM | 0.7033 | 0.6623 | 0.6437 |

**Table 4.** Evaluation for coronary artery semantic segmentation.

| Metrics | Model | LMA | LAD | LCX | D1 | OM1 | RI | Mean |
| --- | --- | --- | --- | --- | --- | --- | --- | --- |
| ACC | RF | 0.9015 | 0.8004 | 0.6211 | 0.5160 | 0.5912 | 0.8059 | 0.6137 |
|  | MLP | 0.8855 | 0.7865 | 0.6751 | 0.5281 | 0.5213 | 0.8093 | 0.6235 |
|  | SVM | 0.8771 | 0.8398 | 0.7461 | 0.6328 | 0.5168 | 0.8235 | 0.7076 |
| IoU | RF | 0.8585 | 0.6039 | 0.5345 | 0.3384 | 0.4187 | 0.7910 | 0.6373 |
|  | MLP | 0.8362 | 0.6370 | 0.5372 | 0.3861 | 0.3876 | 0.7198 | 0.6438 |
|  | SVM | 0.8569 | 0.6796 | 0.6014 | 0.4832 | 0.4037 | 0.8235 | 0.6868 |

According to Table 3, our approach achieved an average accuracy at 0.7033 for coronary artery segment classification. The SVM classifier outperformed other classifiers significantly in ACC, SN and SP. According to Table 4, the proposed approach achieved an accuracy of 0.7076 for pixel classification and 0.6868 for artery semantic segmentation. In Fig. 3, we visualized the results generated at each step for LAO and RAO views.

## 4. Discussion

### 4.1. Performance analysis for artery semantic segmentation

In this paper, a new machine learning based algorithm was applied to perform coronary artery segment classification. The SVM achieved an ACC of 0.7033, a SN of 0.6623 and a SP of 0.6437 among 729 arterial segments in the test set. The handcraft features only contained the low-level image feature, and low-level position features, which limited the model performance. In addition, the number of training samples is not enough, further studies should increase training samples.

For semantic segmentation, the proposed approach achieved a mIoU of 0.6868 and an average pixel classification accuracy of 0.7076. According to the grid search and the performance reported in Table 4, the SVM with a RBF kernel function outperformed the random forest and MLP in ACC and mIoU significantly.

For the proposed approach, errors come from the following three parts: (1) the inaccurate coronary artery segment classification, (2) the coronary artery segments removed due to the limited length, and (3) the mapping method between the centerline and the coronary segment. For the second reason, in our implementation, if the length of the centerline is smaller than a 10-pixel length, then it was removed. However, when evaluating the semantic segmentation performance, the removed artery segments were incorrectly measured, which degraded the model performance. For the third reason, the function for mapping the coronary centerline and segment cannot guarantee a perfect match. In our implementation, we first interpolated the centerline by applying one-dimensional constant interpolation within the sequence of the centerline pixel coordination. Then a perpendicular line was calculated for each interpolated coordinate. If the perpendicular line containing the pixels belonged to the artery in the binary mask, then the pixel was mapped to the centerline. By iterating all the interpolated coordinates in the artery centerline, the arterial pixels were partitioned to the arterial segment. However, the pixels near the bifurcation key points cannot be correctly mapped to the centerline caused by the overlap of the generated perpendicular lines and two-dimensional projection views. These errors may be resolved from different frames and views by operators.

### 4.2. Clinical overview and application

Anatomic delineation of coronary arteries for clinical decisions is best assessed by ICA due to its high spatial resolution and its real time image acquisition. In clinical practice, an automated system for decision support of CAD diagnosis and treatment would require automated anatomical labeling of the coronary artery tree extracted from ICA. By performing the semantic segmentation, this automated process can be achieved with a higher efficiency and reproducibility.

## 5. Conclusion

We propose a machine learning-based approach to classify individual coronary arteries in ICA by incorporating global position and topology information. To precisely extract the entire vascular tree, our previously developed binary segmentation network was employed to generate the arterial binary mask. By generating the centerline of the vascular tree, we identified the key points and converted the centerlines into an artery graph. Each artery segment is represented by an edge in the graph. Finally, we extracted 33 handcraft features of the artery segment and employed a SVM classifier to classify the artery segments and further generated the semantic segmentation results. The proposed approach achieved a mIoU of 0.6868 and an average pixel classification accuracy of 0.7076 for coronary artery semantic segmentation. It shows promise for clinical use to improve efficiency and reproducibility in a cath lab.

## Data Availability

The data is currently unavailable to the public.

## Acknowledgments

This research was supported by a new faculty startup grant from Michigan Technological University Institute of Computing and Cybersystems (PI: Weihua Zhou) and a seed grant from Michigan Technological University Health Research Institute. This research was also supported in part by the National Institutes of Health under award numbers U19AG055373, R01GM109068, R01MH104680, R01MH107354 and by the National Science Foundation NSF under award number 1539067.

